# Active Surveillance for Heartland virus in North Carolina: Clinical and Genomic Epidemiology

**DOI:** 10.64898/2026.02.27.26347100

**Authors:** Diana L. Zychowski, Lauryn Ursery, Sophia Sukkestad, Alaa Ahmed, Dana Giandomenico, Shuntai Zhou, Melissa Miller, Jonathan J. Juliano, Anne Piantadosi, Ross M. Boyce

## Abstract

**Background:** Heartland virus (HRTV) is an emerging tick-borne virus capable of causing severe illness and death. The burden of disease is likely underestimated due to limited seroprevalence studies, lack of commercially available diagnostic tests, and an overlapping clinical syndrome with more commonly diagnosed bacterial diseases such as spotted fever group rickettsiosis or ehrlichiosis.

**Methods:** Active surveillance for Heartland virus disease was conducted at a large academic center from March to September 2024. Enrolled subjects included those who had testing sent for *Ehrlichia* polymerase chain reaction (PCR) along with fever and 2 of the 3 criteria: leukopenia, thrombocytopenia, and/or elevated liver function tests. Specimens with detectable RNA underwent whole genome sequencing and analysis.

**Findings:** Over 800 specimens were received with 53 individuals meeting enrollment criteria. Among these 53, two (3.8%) had detectable HRTV RNA in whole blood during the time of *Ehrlichia* PCR testing. The two cases had disparate clinical manifestations: one with mild disease which was identified in an outpatient setting, while a second case required intensive care unit-level support. Heartland virus genome sequences from the two cases were more similar to viruses from other states than they were to one another.

**Interpretation:** Despite only two prior reported cases of Heartland virus disease in North Carolina, we identified two individuals with acute HRTV viremia. Further surveillance for HRTV disease is necessary to understand the burden of disease and to facilitate further studies of virus pathogenesis and host responses.

**Funding:** Funding for the study was provided by a Creativity Hub Award to Dr. Boyce from the UNC Office of the Vice Chancellor for Research. Dr. Zychowski’s effort was supported by the T32 NIAD grant AI070114.

## Introduction

Heartland virus (HRTV) is an emerging tick-borne virus in the Bandavirus genus (family Phenuiviridae, order Hareavirales) that can cause severe and fatal disease. It is a negative-sense RNA virus with a tripartite genome comprising the small (S), medium (M), and large (L) segments, which encode the nucleoprotein (NP) and nonstructural protein (NSs), the glycoprotein (Gp) subunits Gn/Gc, and the RNA-dependent RNA polymerase (RdRp), respectively. Phylogenetically, HRTV is most closely related to Dabie bandavirus (formerly severe fever with thrombocytopenia syndrome virus), a highly virulent pathogen endemic to Southeast Asia (1). Since the discovery of HRTV in 2009, over 60 cases of heartland virus disease have been reported across 14 states in the midwestern and southeastern United States (1,2) where the primary vector, the lone star tick (*Amblyomma americanum*), is widely distributed (3,4). Clinical manifestations typically include fever, arthralgia, diarrhea, and headache, though cases can progress to critical illness (e.g., sepsis, end organ failure) with documented case fatality rates exceeding 10% (5). Laboratory abnormalities can include thrombocytopenia, leukopenia, elevated aspartate aminotransferase (AST) and/or alanine aminotransferase (ALT) (1,5,6).

The incidence of HRTV disease is almost certainly underreported due to the lack of commercially-available diagnostic tests and the shared clinical syndrome with bacterial tick-borne diseases (TBDs), such as ehrlichiosis and rickettsiosis. Pitfalls in serological bacterial tick-borne testing algorithms, which require a four-fold rise in antibody titers over several weeks, also contribute to the underestimation of TBV incidence. Over 75% of acute serological tests for *Ehrlichia* or *Rickettsia* can lack a paired convalescent sample, and many cases fall into suspected or probable categories rather than a confirmed diagnosis (7). The interpretation of a single, acute, positive titer is challenging, particularly because individuals may have persistent positive serology from past exposure. For example, antibodies to *Ehrlichia* or Spotted fever group *Rickettsia* (SFGR) can be seen in 10-20% of individuals in endemic areas (8). It is therefore conceivable that individuals presenting with acute tick-borne viral disease may incorrectly have their viral symptoms attributed to *Ehrlichia* or *Rickettsia* based upon a single positive serologic titer. This misclassification is particularly troublesome for critically ill patients that expire before confirmatory convalescent testing is performed.

The absence of real time, commercially available nucleic acid assays further complicate the diagnosis of Heartland virus disease. Suspected cases must be referred to the Centers for Disease Control and Prevention (CDC) for HRTV immunoglobulin M (IgM) antibody, reverse transcription-polymerase chain reaction (RT-PCR), and/or plaque reduction neutralization testing (PRNT) (9).

Given the shared spectrum with bacterial tick-borne diseases and the lack of commercially available tests, the true incidence and clinical symptomatology of Heartland virus disease remains unknown. The inability to detect viremia in real-time, resulting in a scarcity of clinical specimens and a lack of published sequences, has also hindered our understanding of TBV pathogenicity and persons at risk for severe disease. Therefore, the goal of this study was to perform active surveillance for HRTV among individuals undergoing testing for ehrlichiosis. Participants with detectable viremia were contacted for detailed clinical and behavioral data collection and invited to participate in peridomestic tick drags to collect ticks for HRTV screening.

## Methods

### Study design and participants

This prospective surveillance study enrolled subjects from the University of North Carolina (UNC) Health System between 3/27/24 and 9/25/24 that underwent testing for ehrlichiosis by polymerase chain reaction (PCR). The UNC network is one of the largest health systems located in North Carolina (NC) and includes over 16 hospitals and oversees over 490,000 emergency room visits annually (10).

Electronic medical record review was performed to collect demographic, laboratory, clinical, treatment, and outcomes data. Enrollment in this study required subjective or objective fever and at least two of the following criteria: leukopenia (white blood cells [WBC] < 4 x 10^9/L), thrombocytopenia (platelets <150 x 10^9/L), or elevated alanine aminotransferase (ALT) and/or aspartate aminotransferase (AST) levels (>45 U/L). The collection of the remnant samples and associated clinical data utilized in the study was approved by the UNC institutional review board (IRB 21-0356).

### Clinical laboratory tests

Spotted fever group *Rickettsia* immunoglobulin G (IgG), *Ehrlichia* IgG, *Ehrlichia* PCR, alpha-gal immunoglobulin E (IgE), and Lyme serology was conducted by McLendon laboratories at UNC hospitals. Specimens with positive Lyme antibodies underwent reflex Western Blot confirmation at Mayo Medical Laboratories. Case status was determined by a combination of clinical and laboratory evidence, as described by the CDC case definitions (11,12). All *Ehrlichia* PCR whole blood specimens were stored at 4°C and were subsequently aliquoted into 400ul amounts with RNA/DNA saver before storage at -80°C.

### Tick-borne viral testing

Whole blood from the *Ehrlichia* PCR remnant sample was tested for HRTV and Bourbon virus (BRBV) RNA using a multiplex RT-PCR assay. Total RNA was extracted from whole blood using Direct-zol RNA Miniprep kit (Zymo Research) and serum or cerebral spinal fluid (CSF) using E.Z.N.A Viral RNA kit (Omega Bio-tek), per manufactures instructions. Primers and probes targeting BRBV nucleoprotein (13) and HRTV NSs (4) were used as previously published. The ubiquitin conjugating enzyme E2 D2 (UBE2D2) and *Amblyomma americanum* tick 16S mitochondrial rRNA (Tick 16S) primer/probe were used as an internal control for human and tick RNA extraction, respectively, as previously published (14,15). Samples were tested in triplicate with positive controls for BRBV (BEI Resources, NIAID, NIH: Genomic RNA from Bourbon Virus, Original, NR-50146), and HRTV via a duplexed DNA oligonucleotide constructed from the HRTV NSs sequence (**Supplemental Table 1**).

### Tick collection and processing

Ticks were collected in November and December of 2024 and were placed in 100% ethanol in transit and subsequently stored at 4°C until they were sorted according to species, sex, and life stage (up to *n*=5 for adult ticks and *n*=25 for nymphs or larvae). Bead bashing tubes were placed into the Disruptor Genie (Scientific Industries, Cat. No. SI-D238) and run at 3000 RPM for 15-minute intervals until ticks were well homogenized. The homogenates were subjected to RNA extraction utilizing Quick-RNA Tissue/Insect Microprep kit (Zymo Research), per the manufacturer’s protocol.

### Sequencing and phylogenetic analysis

Sequencing and phylogenetic analyses were completed as previously described (16). In brief, samples that tested positive for HRTV via RT-PCR underwent full-genome multiplex amplicon library construction using 10, 18, and 36 primer pairs to cover the S, M, and L segments of HRTV, respectively. Libraries were pooled and sequenced on an Illumina MiSeq, and the resulting reads underwent reference-based assembly using viralrecon v2.6.0. All available reference sequences for HRTV S (n=40), M (n=38), and L (n=38) segments were downloaded from GenBank on January 07, 2026. Reference sequences with ≥95% coverage of the coding region for each segment were aligned with the new sequences generated in this study using MAFFT v7.490 in Geneious Prime v2025.2.2, and alignments were trimmed to only include the coding region for each segment. Maximum-likelihood phylogenetic trees were constructed using IQ-TREE v.2.3.5 with 1,000 ultrafast bootstrap replicates. Nucleotide substitution models were selected based on Bayesian Information Criterion (BIC) scoring using ModelFinder Plus (L-segment: GTR+F+I; M-segment: TIM2+F+G4; S-segment: TPM3+I). Trees were formatted, annotated, and rooted on the midpoint in iTOL.

Pairwise and mean nucleotide diversity per state were calculated in RStudio v2025.05.1+513 using Biostrings v2.72.1, ape v5.8-1, and pegas v1.4 packages, using the coding region of each gene and excluding sites with ambiguous data (N). Amino acid substitutions for each coding sequence were identified by manual inspection of translated nucleotide alignments in Geneious Prime.

## Results

A total of 806 samples were collected from 778 unique patients during this 6-month surveillance window (screened cohort), with 53 unique subjects meeting inclusion criteria for tick-borne viral testing (enrolled cohort). The 53 individuals in the enrolled cohort had a median (IQR) age of 51 (39–72) years and included 33 males (62.3%).

Among individuals in the screened cohort (N=778), 380 (48.8%) recalled a tick or insect bite and 246 (31.6%) received at least one dose of doxycycline or equivalent (e.g., amoxicillin, cefuroxime). Most were seen within an outpatient setting (360/778). However, subjects in the enrolled cohort (N=53) were more frequently hospitalized (69.8% vs 20.8% in the screened cohort) and admitted within the medical intensive care unit (ICU) (20.8% vs 3.5% in the screened cohort). Laboratory abnormalities included leukopenia, thrombocytopenia, and elevated AST or ALT in 31/53 (58.5%), 47/53 (88.7%), and 48/53 (90.6%) individuals, respectively, in the enrolled cohort **(Table 1)**. The number of samples received and tested peaked in July **(Figure 1)**.

**Figure 1:**
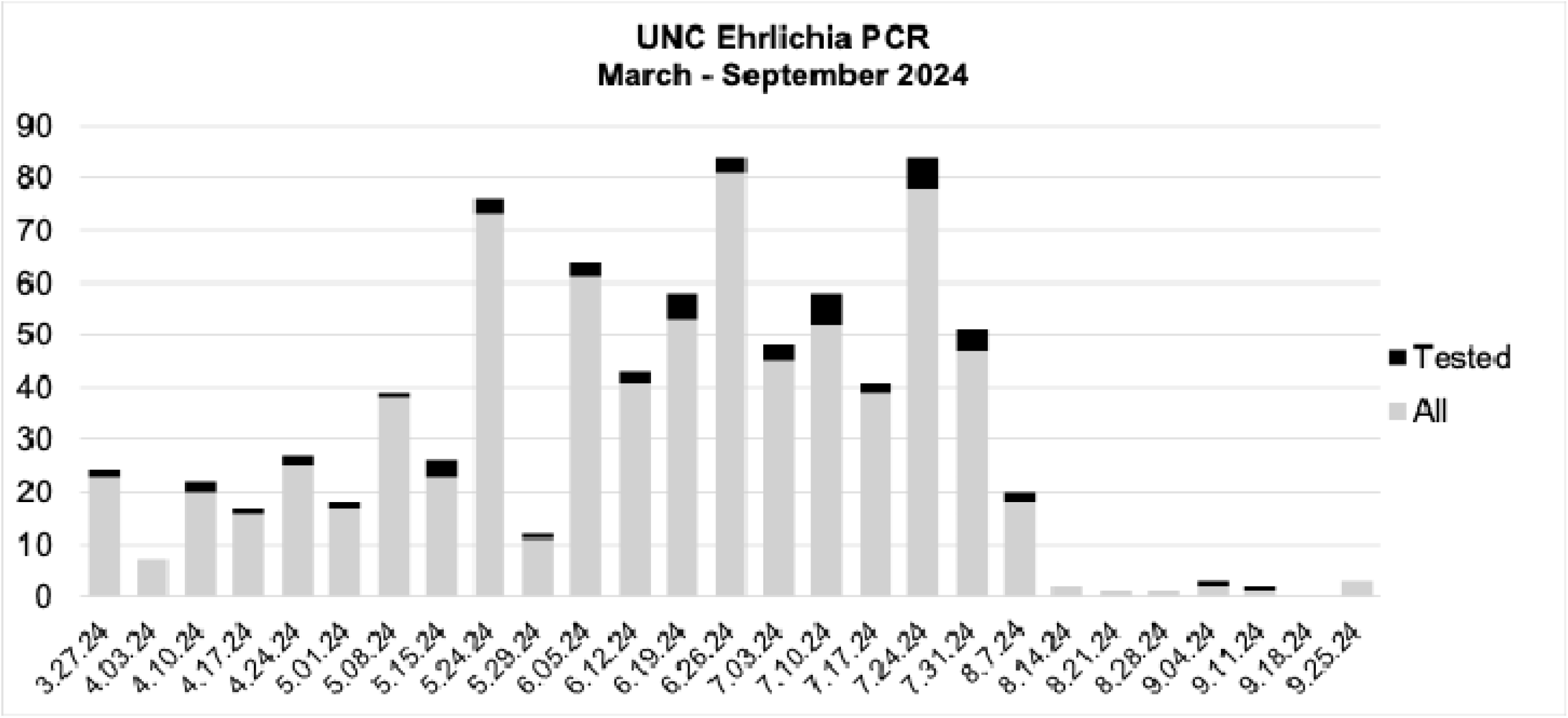
Distribution of collected specimens over time. Figure 1. Distribution of collected remnant *Ehrlichia* polymerase chain reaction (PCR) specimens, including those who underwent TBV testing, between 3/27/24 and 9/25/24.

**Figure 2.**
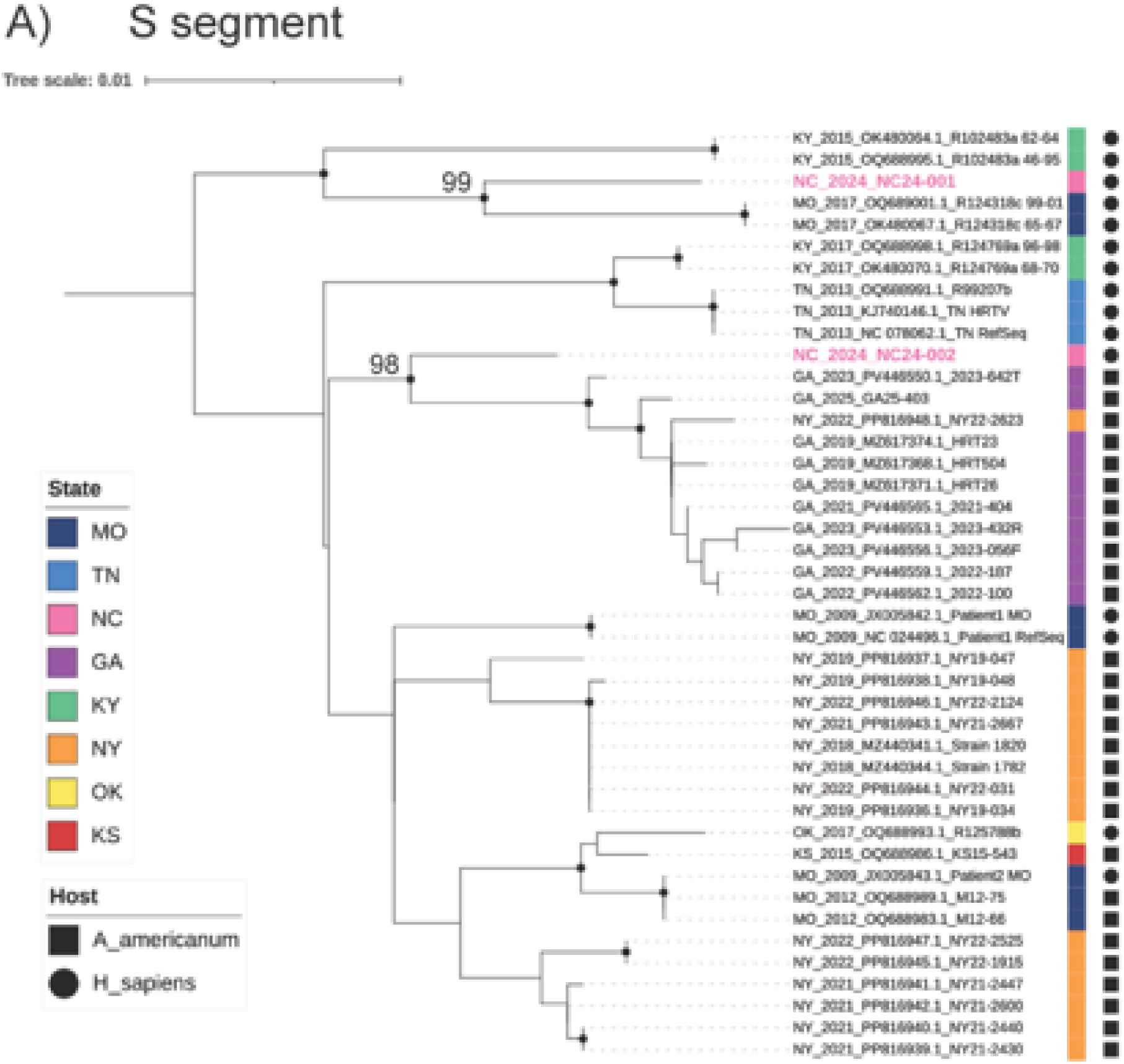
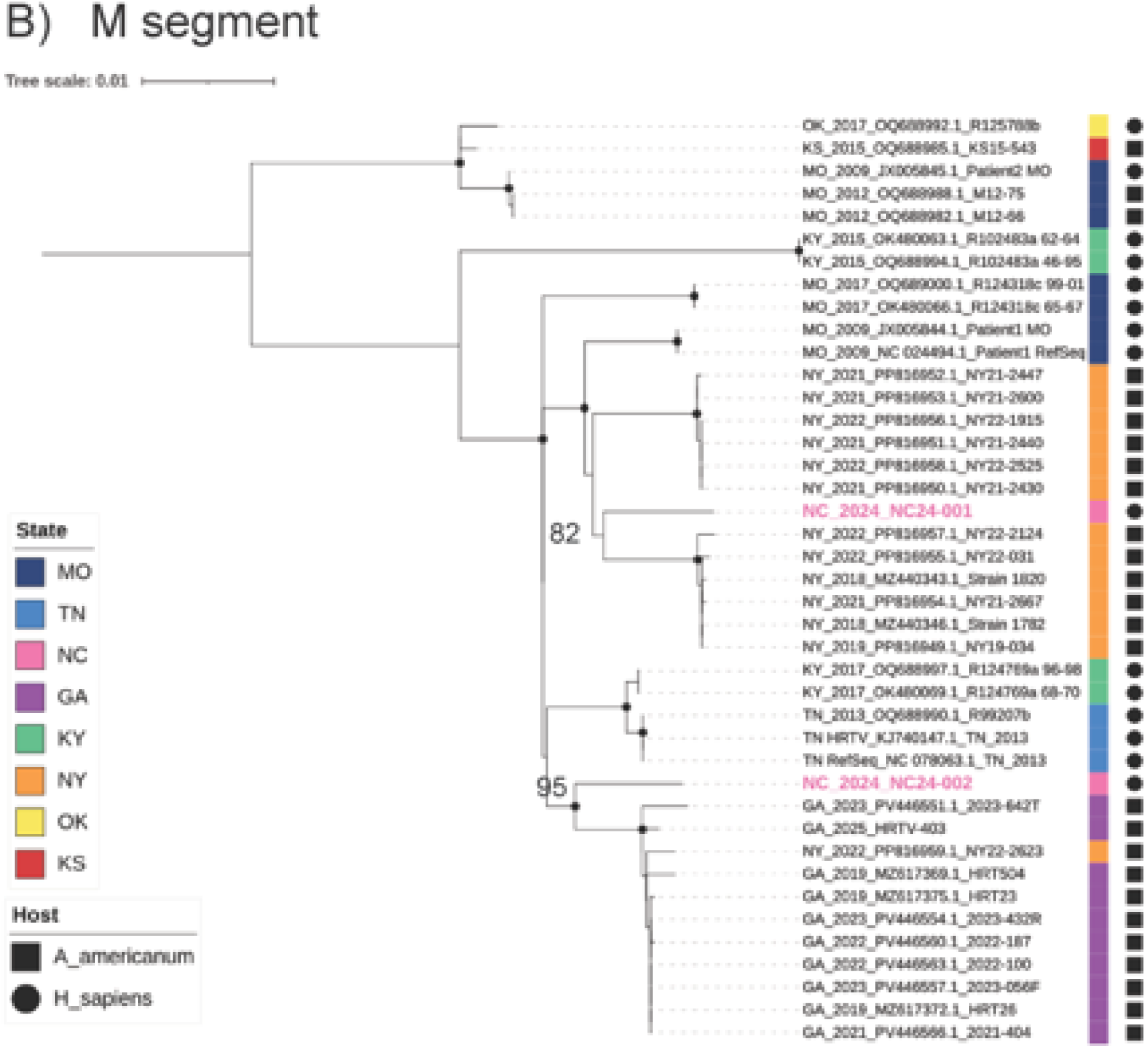
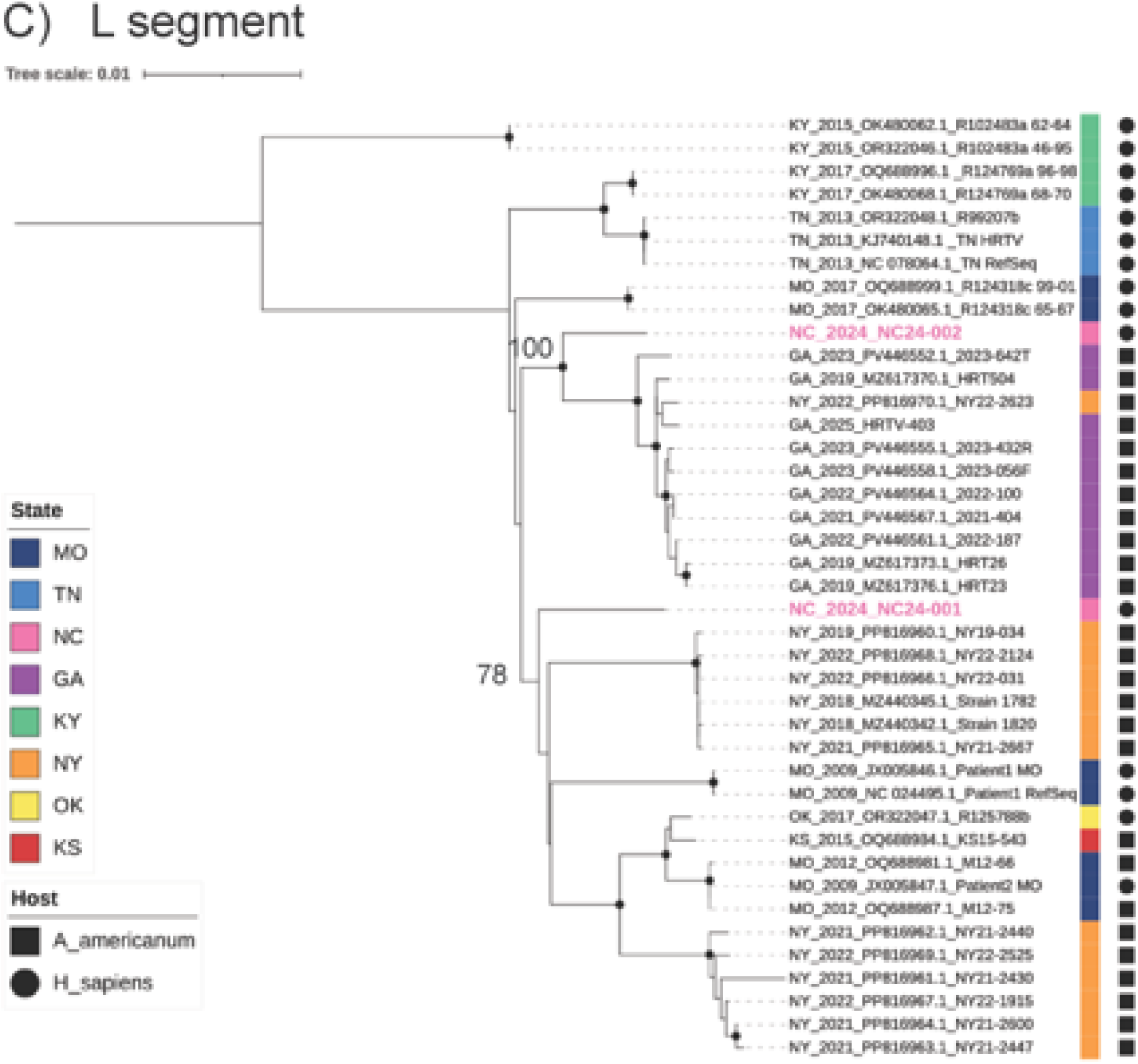
Phylogenetic analysis of HRTV sequences. Figure 2. Likelihood trees are displayed for coding regions of the S segment (A), M segment (B), and L segment (C). Reference sequences from GenBank are named by the strain/isolate name followed by the accession number, state abbreviation for the collection location, and year of collection. Sequences generated in this study are named by sample number, NC, and year collected. NC24-001 and NC24-002 represent sequences from Patients One and Two, respectively. Sequences from ticks are denoted by squares and sequences from human samples by circles. All trees are midpoint rooted. Branches with ultrafast bootstrap values above 0.95 are noted by black dots, and bootstrap values are included for other important nodes.

**Table 1:** Baseline demographic and clinical characteristics of study population.

Abbreviations: IQR, interquartile range; ED, Emergency Department; ICU, intensive care unit; ALT, alanine Aminotransferase; AST, aspartate aminotransferase.
| **Characteristic** | **TBV tested cohort**<br>**No. (%) N=53** | **Screened cohort**<br>**No. (%) N=778** |
| --- | --- | --- |
| Age, median (IQR) | 51 (39,72) | 50 (34,65) |
| Sex |  |  |
| *Male* | 33 (62.3) | 407 (52.3) |
| *Female* | 20 (37.7) | 371 (47.7) |
| Care setting |  |  |
| *Outpatient* | 8 (15.1) | 360 (46.3) |
| *ED/Urgent care* | 8 (15.1) | 254 (32.6) |
| *Inpatient* | 26 (49.1) | 135 (17.4) |
| *ICU* | 11 (20.8) | 27 (3.5) |
| Leukopenia | 31 (58.5) | 56/519 (10.8) |
| Thrombocytopenia | 47 (88.7) | 85/518 (16.4) |
| Elevated ALT/AST | 48 (90.6) | 140/447 (29.4) |
| Tick/Insect bite reported |  |  |
| *Yes* | 10 (18.9) | 380 (48.8) |
| *No* | 28 (52.8) | 195 (25.1) |
| *Not indicated* | 15 (28.3) | 203 (26.1) |
| Alternative diagnosis found |  |  |
| *Yes* | 24 (45.3) | 246 (31.6) |
| *No* | 29 (54.7) | 517 (66.5) |
| *Unknown* | 0 | 15 (2.1) |

Over 95% of individuals in the screened cohort had acute *Ehrlichia* and SFGR serological testing performed, and 340/778 (43.7%) had at least one positive acute or convalescent IgG antibody for *Ehrlichia* and/or SFGR. Ultimately, 83% did not meet any case definition for SFGR or ehrlichiosis. Seven individuals met criteria for confirmed ehrlichiosis or SFGR, four of whom were in the enrolled cohort (N=4/53, 7.5%). Many individuals, 15/53 (28.3%) in the enrolled cohort, and 88/778 (11.3%) in the screened cohort, met criteria for a probable case of ehrlichiosis **(Table 2, Supplemental figure 2)**. Independent of tick-borne disease testing, an alternative diagnosis (e.g., pyelonephritis, bacteremia, systemic lupus erythematosus) was found in nearly two thirds of individuals (246/778 vs 24/53).

**Table 2:** Tick-borne disease testing outcomes.

Tick-borne laboratory test results and case ascertainment. Abbreviations: HRTV, Heartland virus; IgG, immunoglobulin G; SFGR, Spotted fever group *Rickettsia*; Ab, antibody; WB, Western Blot; Alpha-gal IgE, galactose-alpha-1,3-galactose immunoglobulin E.
| **TBD Testing** | **TBV tested cohort**<br>**No. (%) N=53** | **Screened cohort**<br>**No. (%) N=778** |
| --- | --- | --- |
| HRTV RNA + | 2-4 (3.8-7.5) | 2 (0.3) |
| *Ehrlichia* PCR + |  |  |
| *Positive* | 1 (1.9) | 2 (0.3) |
| *Negative* | 52 (98.1) | 776 (99.7) |
| *Ehrlichia* IgG Acute |  |  |
| $\geq 1:128$ | 15 (28.3) | 239 (30.7) |
| $< 1:128$ | 38 (71.7) | 514 (66.1) |
| *Unknown/Not done* | 0 (0.0) | 25 (3.2) |
| *4x rise\** | 1 (1.9) | 2 (0.3) |
| SFGR IgG Acute |  |  |
| $\geq 1:128$ | 8 (15.1) | 158 (20.3) |
| $< 1:128$ | 45 (84.9) | 606 (77.9) |
| Unknown/Not done | 0 (0.0) | 14 (1.8) |
| 4x rise** | 2 (3.8) | 3 (0.4) |
| Lyme |  |  |
| Lyme Ab+, WB IgM + | 0 | 31/760 (4.1) |
| Lyme Ab+, WB IgG + | 0 | 3/760 (0.4) |
| Lyme Ab+, WB IgM and IgG+ | 0 | 2/760 (0.3) |
| Alpha-gal IgE $\geq 0.10$ kAU/L | 0 | 5/43 (11.6) |
| Case definition |  |  |
| Confirmed ehrlichiosis | 2 (3.8) | 4 (0.5) |
| Confirmed SFGR | 2 (3.8) | 3 (0.4) |
| Probable ehrlichiosis | 15 (28.3) | 88 (11.3) |
| Probable SFGR | 7 (13.2) | 48 (6.2) |
| Suspected ehrlichiosis | 0 (0.0) | 7 (0.9) |
| Suspected SFGR | 0 (0.0) | 1 (0.1) |
| Not a case | 31 (58.5) | 646 (83.0) |

\*Only 11/53 and 70/778 had paired convalescent *Ehrlichia* testing
\*\*Only 13/53 and 64/778 had paired convalescent SFGR testing

Two individuals had detectable HRTV RNA via RT-PCR, one of whom had positive HRTV IgM. None had detectable BRBV RNA **(Table 2**). Their courses are as follows:

### Clinical cases

#### Patient One

A woman in her fifties with a history of hypertension, anxiety and depression presented to the Emergency Department (ED) in May 2024 with complaints of generalized malaise, diarrhea, myalgias, arthralgias, sore throat, and fever over the preceding two weeks. She experienced a tick bite but noted an attachment for less than 24 hours.

Prior to her ED presentation, she was evaluated by an outside provider who raised concern for a possible tick-borne infection; however, no diagnostic testing was performed, and she was instructed to take empiric doxycycline.

Upon presentation to the ED she was hypertensive (139/76), febrile (100.6°F), and non-toxic appearing. Notable lab abnormalities included leukopenia (WBC 2.8 x 10^9/L), thrombocytopenia (platelets 137x10^9/L), hyponatremia (Na 133 mmol/L), and elevated liver transaminases (AST 61 U/L, ALT 143 U/L, and alkaline phosphatase 236 U/L). Antibodies to *Ehrlichia*, SFGR, and Lyme were negative. *Ehrlichia* PCR was negative. Viral respiratory testing was negative. She was sent home with a 14-day prescription for doxycycline 100mg twice daily. The subject was contacted following her positive HRTV RNA test result, but she declined further participation.

#### Patient Two

A man in his seventies with poorly controlled type two diabetes mellitus (hemoglobin A1c 9.0%) presented to urgent care in late July 2024 with four days of objective fever, fatigue, myalgias, nausea and chills. He denied any respiratory symptoms and tested negative for COVID-19. He reported multiple attached ticks nearly 12 days prior, raising concern for a tick-borne illness. Antibody testing to *Ehrlichia*, SFGR, and Lyme was sent, though routine bloodwork was not ordered. *Ehrlichia* PCR was negative. He had no rash and was non-toxic appearing. He was sent home with a 10-day prescription for doxycycline 100mg twice daily and was advised to follow up with his primary care provider.

Over the next 96 hours, he returned to the ED two additional times with new and progressive symptoms despite doxycycline use, including increased somnolence, headache, blurred vision and vertigo. He was hypertensive, afebrile, and had no gross neurological deficits. Laboratory testing revealed leukopenia, thrombocytopenia, hyponatremia, and elevated liver transaminases. At this time, his prior tick-borne serological tests revealed reactive antibodies to *Ehrlichia* ≥1:128. He was discharged home with suspicion for ehrlichiosis.

Despite doxycycline, his mentation deteriorated, and he returned to the ED nearly five days after he originally sought care. On arrival, he was febrile and experienced a witnessed seizure. He was admitted to the medical ICU where he was treated with broad spectrum antibiotics including doxycycline, acyclovir, and antiepileptics. Non-contrasted computed tomography of the head and magnetic resonance imaging of the brain without and with contrast material was negative for acute intracranial pathology.

A lumbar puncture was performed and CSF analysis of tubes 1 & 4 demonstrated: 157-1 red blood cells, 4-8 nucleated cells with >90% lymphocytes, protein 249mg/dL, and glucose 140mg/dL. The CSF was negative for herpesviruses and enterovirus via PCR. Bacterial CSF cultures were without growth.

Serum was sent to the CDC, nearly three weeks after the suspected tick exposure, and eventually returned positive for HRTV IgM and HRTV RNA but was negative for HRTV neutralizing antibodies via PRNT. Serum was negative for BRBV RNA and neutralizing antibodies. He had detectable HRTV RNA via our in-house RT-PCR multiplex assay for seven days during his hospitalization, which was nearly 18-25 days following the suspected exposure. His CSF was negative for HRTV RNA via RT-PCR. Cell culture inoculated with his CSF demonstrated a cytopathic effect suggestive of viral infectivity; however, subsequent attempts at viral titration failed to detect quantifiable virus.

Eventually, his mentation improved, and he was discharged to a skilled nursing facility. When seen at outpatient follow up, he reported overall improvement in neurologic function and had no significant deficits outside of intermittent brain fog.

### Tick RT-PCR

Tick drags at the area of exposure for Patient Two were performed in November 2024, and again in April and June 2025 (**Supplemental table 2**). Over 500 ticks were collected, including Asian longhorned (*Haemaphysalis longicornis*) nymphs. None tested positive for HRTV or BRBV RNA.

### Sequence analysis

Phylogenetic analysis of the HRTV S, M, and L genome segments showed distinct clustering between the sequences from Patient One and Patient Two. The HRTV sequences from Patient Two clustered with, and just outside, a clade of HRTV sequences from ticks in Georgia collected from 2019-2025, with at least 95% ultrafast bootstrap support in each segment (**Figure 3**). HRTV sequences from Patient One clustered separately from Patient Two in all genome segments, but in different locations on the tree (**Figure 3**). In the S segment, the Patient One sequence clustered with sequences derived from patients in Missouri in 2017, with 99% ultrafast bootstrap support. In the M and L segments, the Patient One sequences clustered with sequences derived from New York and Midwest states, but with only ∼80% ultrafast bootstrap support.

At the nucleotide level, sequences from Patients One and Two were 3.0% different in the NP gene, 3.1% different in the NSs gene, 2.2% different in the Gp gene, and 1.5% different in the RdRp gene (**Table 3**). This level of pairwise nucleotide diversity was similar to some states (KY, MO) but not others (GA, TN). Interestingly, pairwise nucleotide diversity seemed to be higher in states from which most sequences were derived from human infection (KY, NC, MO) compared to states in which most sequences were derived from ticks (NY, GA). Sequences from Patients One and Two were fairly conserved at the amino acid level, although there was 1 amino acid difference in the NP protein, 3 in NSs, 9 in Gp, and 6 in RdRp (**Table 4**). None of these substitutions were at positions that have previously been implicated in pathogenesis of HRTV or Dabie bandavirus (17,18). More broadly, we did not observe any amino acid substitutions shared more commonly by HRTV sequences from humans compared to ticks.

**Table 3.**
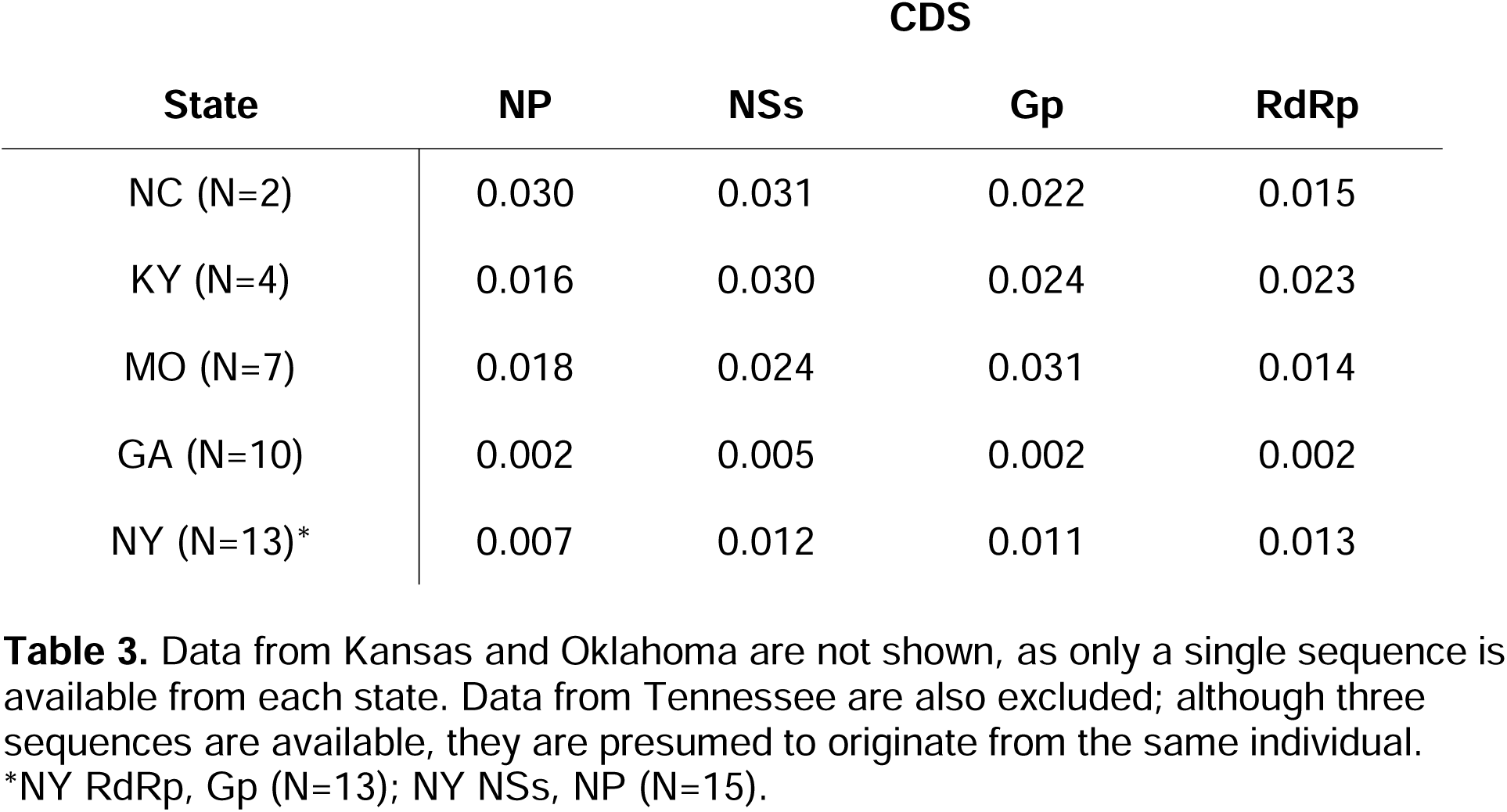
Mean pairwise nucleotide diversity (π) for sequences from each state, by coding region (CDS).

Data from Kansas and Oklahoma are not shown, as only a single sequence is available from each state. Data from Tennessee are also excluded; although three sequences are available, they are presumed to originate from the same individual.
| State | CDS |  |  |  |
| --- | --- | --- | --- | --- |
|  | NP | NSs | Gp | RdRp |
| NC (N=2) | 0.030 | 0.031 | 0.022 | 0.015 |
| KY (N=4) | 0.016 | 0.030 | 0.024 | 0.023 |
| MO (N=7) | 0.018 | 0.024 | 0.031 | 0.014 |
| GA (N=10) | 0.002 | 0.005 | 0.002 | 0.002 |
| NY (N=13)* | 0.007 | 0.012 | 0.011 | 0.013 |

\*NY RdRp, Gp (N=13); NY NSs, NP (N=15).

**Table 4.** Amino acid substitutions per coding region.

Amino acid changes are noted compared to reference sequences NC\_024496.1 (S segment; NP and NSs coding regions), NC\_024494.1 (M segment; Gp coding region), and NC\_024495.1 (L segment; RdRp coding region).
| Sample ID | NP | NSs | Gp | RdRp |
| --- | --- | --- | --- | --- |
| NC24-001 | - | **S31N**;<br>**G237E**;<br>I273V | **S112N** *; I462V; V526I;<br>V613M | T157S; **K279E** *; S302N; **N458S** *;<br>T783A; K856R*; **A1053T** *; E1207D;<br>**F1299S** *; T1340I; T1653A; R2037E;<br>P2038R; G2040C; R2041G |
| NC24-002 | F39L | I273V | **F85Y** *; **M298L**; **P327A** *;<br>I462V; **F487S**; **T560S** *;<br>V613M; **R964K** *; **A1018T** * | T157S; S302N; T783A; E1207D;<br>T1340I; T1653A; **A1938S**; R2037E;<br>P2038R; G2040C; R2041G |

## Discussion

Tick-borne viral testing of 53 individuals undergoing PCR testing for ehrlichiosis identified two patients with detectable HRTV RNA, thereby doubling the number of previously reported HRTV disease cases in North Carolina (19).

The two cases illustrated a spectrum of severity, from persistent flu-like illness in an immunocompetent patient to ICU-level care for HRTV encephalitis in a patient with poorly controlled diabetes. The disparate clinical manifestations seen in this study are not surprising, as there are likely to be asymptomatic cases or more subtle manifestations of HRTV infection, particularly in immunocompetent hosts. For example, a 2013 seroprevalence study identified HRTV IgG and neutralizing antibodies in 2.5% (12/487) and 1.4% (7/487) of Missouri blood donors, respectively (20). Similarly, we previously demonstrated the presence of BRBV-specific neutralizing antibodies in 4/518 (0.77%) individuals from a NC convenience cohort that had no prior clinical syndrome compatible with a TBV (21). It is therefore conceivable that mild cases of TBV disease, whose symptoms may overlap with those of other viral or vector-borne diseases, are rarely diagnosed and consequently seldom reported. Nonlethal cases, along with a lack of TBV seroprevalence studies, limit accurate disease estimation.

Active surveillance, as demonstrated in this study, is needed to define the full clinical spectrum of HRTV disease and to identify viral genomic and host immunologic factors associated with virulence. Prior studies have shown that impaired interferon responses enhance HRTV pathogenicity and that the HRTV NSs functions as a virulence factor by antagonizing interferon induction and signaling pathways (22–24). However, only two amino acid differences – one nonconservative – were observed in NSs between our cases, and given limited in vitro data, their functional impact remains uncertain.

Our study generated the first complete HRTV genome sequences from NC, and notably, these viruses were phylogenetically divergent. Similar genetic diversity has been observed in other states with multiple HRTV cases, however, tick-derived sequences tend to cluster geographically. We also observed segment-specific clustering in Patient One, raising the possibility of reassortment, however the trees were not statistically robust to formally assess this. These patterns may reflect human infection acquired across diverse locations; long-range dispersal of HRTV-infected ticks, and/or viral evolution following human infection. Broadly, these findings highlight substantial gaps in our understanding of HRTV genetic diversity, evolution, and transmission.

The absence of HRTV RNA from pooled ticks was unsurprising given the low minimum infection rate 0.46-9.46/1000 for lone star ticks (25,26). However, the presence of Asian longhorned ticks (*Haemaphysalis longicornis*) was notable. This recently established species is the primary vector of Dabie bandavirus found in Asia (27,28), and has been shown experimentally to acquire and transmit HRTV, consistent with the phylogenetic relatedness of these viruses (29). Although HRTV has not yet been detected in environmentally-collected longhorned ticks, their ability to transmit HRTV and other pathogens (e.g., borrelia, anaplasma, and bartonella), along with rapid population establishment through parthenogenesis, underscores the need for close surveillance (30).

As our study shows, there is also a need for increased surveillance for human cases of HRTV and improved clinical diagnostic strategies. Increased use of TBV RT-PCR assays can provide expedited results which can i) help healthcare providers avoid unnecessary testing and treatment and provide anticipatory guidance ii) help distinguish TBVs from endemic bacterial tick-borne diseases and iii) facilitate the collection of biospecimens for further studies. The prolonged duration of HRTV viremia observed in this study (18–25 days post–tick bite) and in prior reports (8–14 days post–illness onset, and up to 23 days in an immunocompromised individual)(5) also supports a relatively broad window for diagnostic testing. These data support TBV testing in areas endemic for lone star or Asian longhorned ticks, with a stepwise approach in resource-limited settings that prioritizes patients who fail doxycycline within 48–72 hours or meet defined clinical criteria.

Beyond timely TBV testing, improved diagnostic stewardship for bacterial TBDs is needed. In this study, completion of serological convalescent testing was poor: only 9.0% (N=70/778) and 8.2% (N=64/778) of screened individuals had completed paired testing for *Ehrlichia* and *Rickettsia*, respectively. Improving provider education on tick-borne testing algorithms and expanding telemedicine in rural areas may improve testing completeness and case ascertainment (7). Completion of paired serological testing is particularly important in highly endemic areas which may have a high background prevalence of TBDs antibodies. In our screened cohort, 239 (30.7%) and 158 (20.3%) individuals had detectable IgG antibodies to *Ehrlichia* and SFGR, respectively. Notably, 31.6% (N=246/778) of screened individuals were later diagnosed with a non-tick-borne condition, yet 30 of the 246 individuals met the probable or suspect case definition for ehrlichiosis and/or rickettsiosis, highlighting the limited specificity in traditional case definitions. Conversely, there were five individuals within our tested cohort who reported a tick bite, had a clinical syndrome consistent with a TBD, and had no detectable bacterial antibodies, suggesting an alternative pathogen (e.g., Powassan virus, *Borrelia)* or false-negative testing. Collectively, these findings underscore the limited specificity of single antibody titers to predict acute bacterial disease and the need for novel diagnostic approaches.

This study had many strengths. In partnership with an invested clinical laboratory, we were able to acquire specimens, perform testing, and provide results quickly. As a result, we were able to contact cases to obtain complementary clinical and behavioral data, perform tick drags, and collect specimens for sequencing and biobanking. However, our results likely underestimate incident TBV infections as it is possible that we missed individuals with recovered HRTV infection or low-level viremia. In addition, given that viral testing was only conducted on those with specific clinical and laboratory criteria, cases of HRTV disease may have a wider spectrum of symptomatology and objective laboratory findings than described.

## Conclusion

Despite only two prior reported cases of Heartland virus disease in North Carolina, we identified two individuals with acute HRTV viremia during a 6-month window of active surveillance. Our findings substantially expand our understanding of clinical HRTV disease and provide the foundation for which future genomic and immunological studies can explore the pathogenesis of emerging TBVs. Investment in surveillance systems and diagnostic testing is needed to improve our understanding of disease burden and facilitate therapeutic development for emerging pathogens.

## Data Availability

All data produced in the present study are available upon reasonable request to the authors

## Acknowledgments

### Potential Conflicts of Interests

All authors report no financial relationships with any organizations that might have an interest in the submitted work in the previous three years except that noted in the funding section; no other relationships or activities that could appear to have influenced the submitted work.

### Funding

Funding for the study was provided by a Creativity Hub Award to RMB from the UNC Office of the Vice Chancellor for Research. Dr. Zychowski’s effort was supported by the T32 NIAD grant AI070114. This study was supported in part by the Emory Integrated Genomics Core (EIGC), which is subsidized by the Emory University School of Medicine and is one of the Emory Integrated Core Facilities.

Role of the funder/sponsor(s): The funder(s) had no role in the design and conduct of the study; collection, management, analysis, and interpretation of the data; preparation, review, or approval of the manuscript; and decision to submit the manuscript for publication.

### Author Contributions

Diana Zychowski is a physician-scientist at Hennepin Healthcare Research Institute, and recent Infectious Disease Fellow at the University of North Carolina at Chapel Hill. Her primary research focus is on the intersection of tick-borne infections, public health surveillance, and population health sciences. Dr. Zychowski has full access to all the data in the study and take responsibility for the integrity of the data and the accuracy of the data analysis.

Study conception and design: RMB, DLZ. Funding: RMB, DLZ. Laboratory testing: DLZ, SS, AA, AP. First draft of manuscript: DLZ. Revisions: All.

## Acknowledgments

We would like to thank McLendon Labs for testing and coordination with specimen acquisition. Dr. Helen Lazear and her team for cell culture work. Brooke Merdjane, Ester Lee, and Molly Munoz for specimen processing. As well as the CDC for testing confirmation.

## Notes

### Competing Interest Statement

The authors have declared no competing interest.

### Author Declarations

IRB of the University of North Carolina at Chapel Hill gave ethical approval for this work. IRB 21-0356.

